# Self-Assessment of amyotrophic lateral sclerosis functional rating scale on the patient’s smartphone proves to be non-inferior to clinic data capture

**DOI:** 10.1101/2024.12.03.24318381

**Authors:** Laura Steinfurth, Torsten Grehl, Ute Weyen, Dagmar Kettemann, Robert Steinbach, Annekathrin Rödiger, Julian Großkreutz, Susanne Petri, Matthias Boentert, Patrick Weydt, Sarah Bernsen, Bertram Walter, René Günther, Paul Lingor, Jan Christoph Koch, Petra Baum, Jochen H. Weishaupt, Johannes Dorst, Yasemin Koc, Isabell Cordts, Maximilian Vidovic, Jenny Norden, Peggy Schumann, Péter Körtvélyessy, Susanne Spittel, Christoph Münch, André Maier, Thomas Meyer

## Abstract

**Objective:** To investigate self-assessment of the amyotrophic lateral sclerosis functional rating scale–revised (ALSFRS-R) using the patient’s smartphone and to analyze non-inferiority to clinic assessment.

**Methods:** In an observational study, ALSFRS-R data being remotely collected on a mobile application (App-ALSFRS-R) were compared to ALSFRS-R captured during clinic visits (clinic-ALSFRS-R). ALS progression rate (ALSPR) – as calculated by the monthly decline of ALSFRS-R – and its intrasubject variability (ALSPR-ISV) between ratings were used to compare both cohorts. To investigate non-inferiority of App-ALSFRS-R data, a non-inferiority margin was determined.

**Results:** 691 ALS patients using the ALS-App, and 1895 patients with clinic assessments were included. Clinical characteristics for the App-ALSFRS-R and clinic-ALSFRS-R cohorts were as follows: Mean age 60.45 (SD 10.43) and 63.69 (SD 11.30) years (p<0.001), disease duration 38.7 (SD 37.68) and 56.75 (SD 54.34) months (p<0.001) and ALSPR 0.72 and 0.59 (p<0.001), respectively. A paired sample analysis of ALSPR-ISV was applicable for 398 patients with clinic as well as app assessments and did not show a significant difference (IQR 0.12 [CI 0.11, 0.14] vs 0.12 [CI 0.11, 0.14], p=0.24; Coheńs d =0.06). CI of IQR for App-ALSFRS-R was below the predefined non-inferiority margin of 0.15 IQR, demonstrating non-inferiority.

**Conclusions:** Patients using a mobile application for remote digital self-assessment of the ALSFRS-R revealed younger age, earlier disease course and faster ALS progression. The finding of non-inferiority of App-ALSFRS-R assessments underscores, that data collection using the ALS-App on the patient’s smartphone can serve as additional source of ALSFRS-R in ALS research and clinical practice.

## INTRODUCTION

The ALS Functional Rating Scale in its revised version (ALSFRS-R) is a severity score reflecting the course of ALS (1,2). The 12-item scale is disease-specific and was designed to assess bulbar symptoms, limb and trunk functions, respiratory symptoms and the need of ALS-related interventions such as percutaneous endoscopic gastrostomy, non-invasive ventilation or tracheostomy with invasive ventilation (3). The scale was primarily developed as an outcome parameter in clinical trials but evolved to the most widely applied rating scale in both clinical practice and ALS research (4).

The ALSFRS-R is commonly captured during clinic visits. However, clinic consultations can be burdensome for people with ALS, especially with progressing impairment. As the ALSFRS-R does not rely on physical examination, remote assessment via telephone as well as online was proposed (4–6). This aims to reduce the efforts of clinic assessment and to complement data gaps between clinic visits (7,8). The feasibility of the ALSFRS-R for self-rating in terms of a patient-reported outcome, paved the way for its remote digital assessment using online platforms and apps (4,9–12). Nowadays self-assessments and online questionnaires are becoming part of standard practice, using adapted versions of the ALSFRS-R. Furthermore, remote self-assessment may increase the efficiency of clinical studies if the rating of ALSFRS-R is moved to digital data capture (13,14). To support the concept of remote digital assessment, in 2022 a German consensus group developed an annotated German and English version of the ALSFRS-R scale that is self-explanatory and unambiguous (ALSFRS-R-SE) (15).

Despite its limitations (16–18), the ALSFRS-R serves as a meaningful clinical decision-making criterion in ALS care and established outcome parameter in clinical trials. Furthermore, the ALSFRS-R is basis for calculating the ALS progression rate (ALSPR), that puts the total-score of the ALSFRS in relation to the disease duration. The ALSPR is recognized as an independent predictor of survival and was correlated with ALS phenotypes and the biomarker neurofilament light chain (19–22). In clinical trials, ALSPR was applied for patient selection, as well as stratification (21–24). Only recently, ALSPR – as assessed in clinic and remotely – was applied to quantify the treatment response to tofersen (25,26).

Given the increasing use of digital platforms and mobile applications, in particular the introduction of the “ALS-App” in Germany, Austria, and Switzerland, remotely captured ALSFRS-R data have become increasingly available. At the same time, there are uncertainties about the extent to which the data collected remotely is comparable with clinic assessments (27). Here, we report the investigation of remote digital assessment of ALSFRS-R by using a mobile application (ALS-App). The aims of the present study were 1) to assign and evaluate demographic and clinical characteristics to the cohorts of clinic and app assessments, 2) to compare the intrasubject variability of ALSPR of clinic and app data and 3) to investigate if non-inferiority of remote digital assessment using the ALS-App compared to clinic assessment of ALSPR can be proven.

## METHODS

### Study design

The observational study was conducted as a prospective, multicentre cohort study. The investigation was reported according to the STROBE criteria (28,29). The study was conducted from May 2020 until April 2024.

### Participants

The participants met the following inclusion criteria: 1) diagnosis of ALS according to the Gold Coast criteria (28), (2) consent to electronic data capture using the research platform “APST”; 3) capture of at least two assessments of ALSFRS-R.

### Setting

#### App assessment of ALSFRS-R using the ALS-App (App-ALSFRS-R)

Patients were offered a remote digital assessment of the ALSFRS-R on a mobile application (ALS-App), which may be used on smartphones or tablet devices and was available for iOS and Android devices (https://www.ambulanzpartner.de/als-app/). After obtaining informed consent, patients received an activation link for the digital data capture. For technical support, a telephone service and email contact were provided. All patients were requested to digitally complete the ALSFRS-R at least once a month. An email reminder was sent accordingly.

#### Clinic assessment of ALSFRS-R (clinic-ALSFRS-R)

16 multidisciplinary ALS centres in Germany and Austria participated in this study and provided ALSFRS-R data and clinical data, obtained during the regular visits. Those served as source data for this study (secondary use of existing data for research purposes). The evaluators consisted of neurologists, study nurses and coordinators, who were trained in ALSFRS-R assessment.

#### Description of cohorts

All participants who met the inclusion criteria formed the total study cohort. The “clinic-ALS-FRS-R cohort” included patients with ALSFRS-R assessments during clinic visits. The “App-ALSFRS-R cohort” encompassed participants who performed remote digital assessment of ALSFRS-R using the ALS-App. The “combined ALSFRS-R cohort” included patients who provided at least two assessments in both settings, i.e. clinic and app assessment, at any time during the observation period **(Figure 1).**

**Figure 1.**
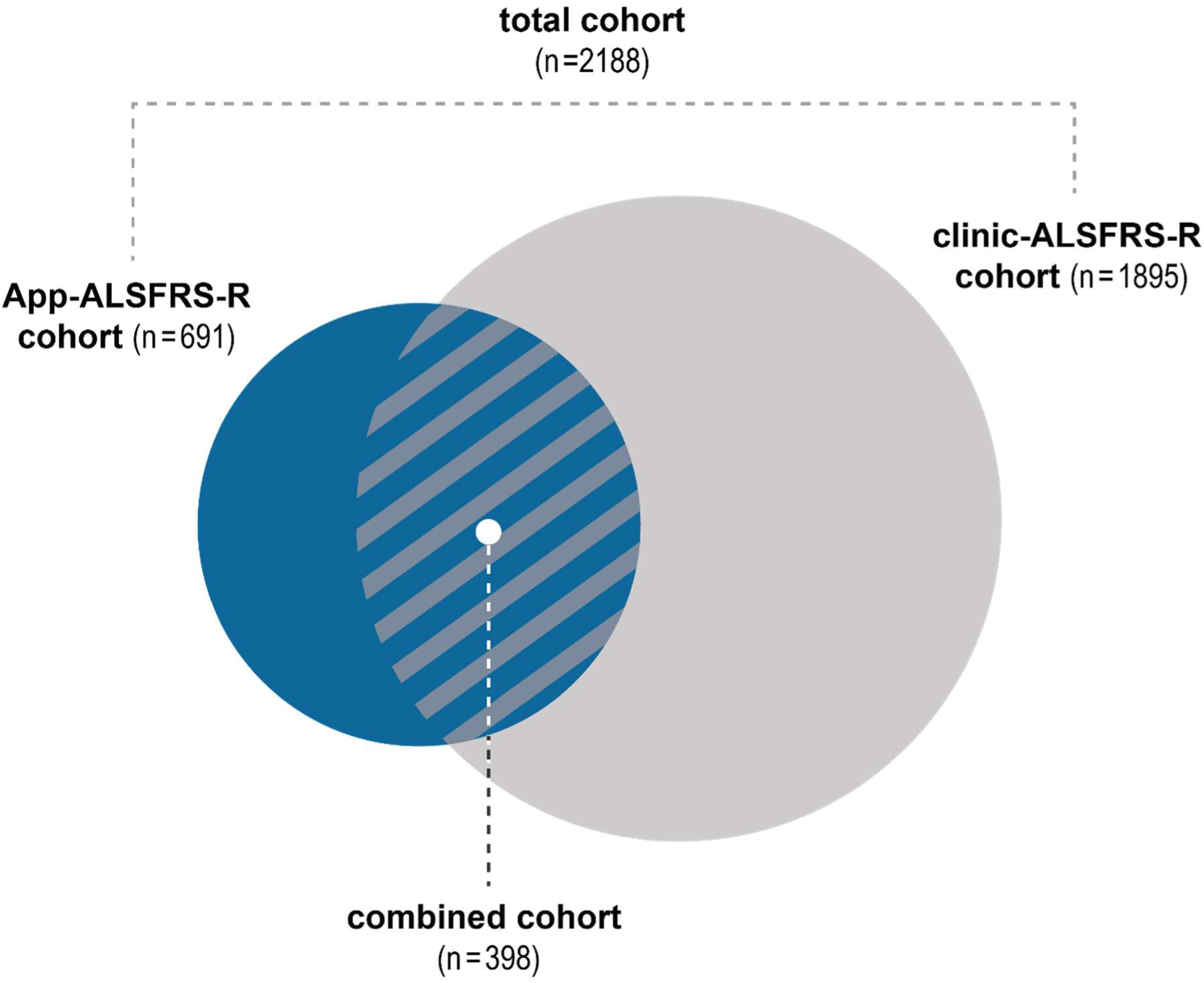
Overview of studied cohorts: remote digital assessment of the ALSFRS-R using the “ALS App” (App-ALSFRS-R) and the assessment during clinic visits (clinic-ALSFRS-R) were investigated. A sub-cohort performed both, ALSFRS-R assessment during clinic visits and remote rating via the “ALS App” (combined cohort). n=number of patients, ALSFRS-R=Amyotrophic Lateral Sclerosis Functional Rating Scale revised.

#### Protocol approvals and registrations

The study protocol was approved by the Medical Ethics Committee of Charité – Universitätsmedizin Berlin, Germany under number EA1/219/15. A signed patient information and informed consent form was obtained from all the participating patients.

### Variables

#### Demographic and clinical characteristics

The following demographic and clinical characteristics were collected: age, sex, onset of symptoms **(Table 1)**.

**Table 1:**
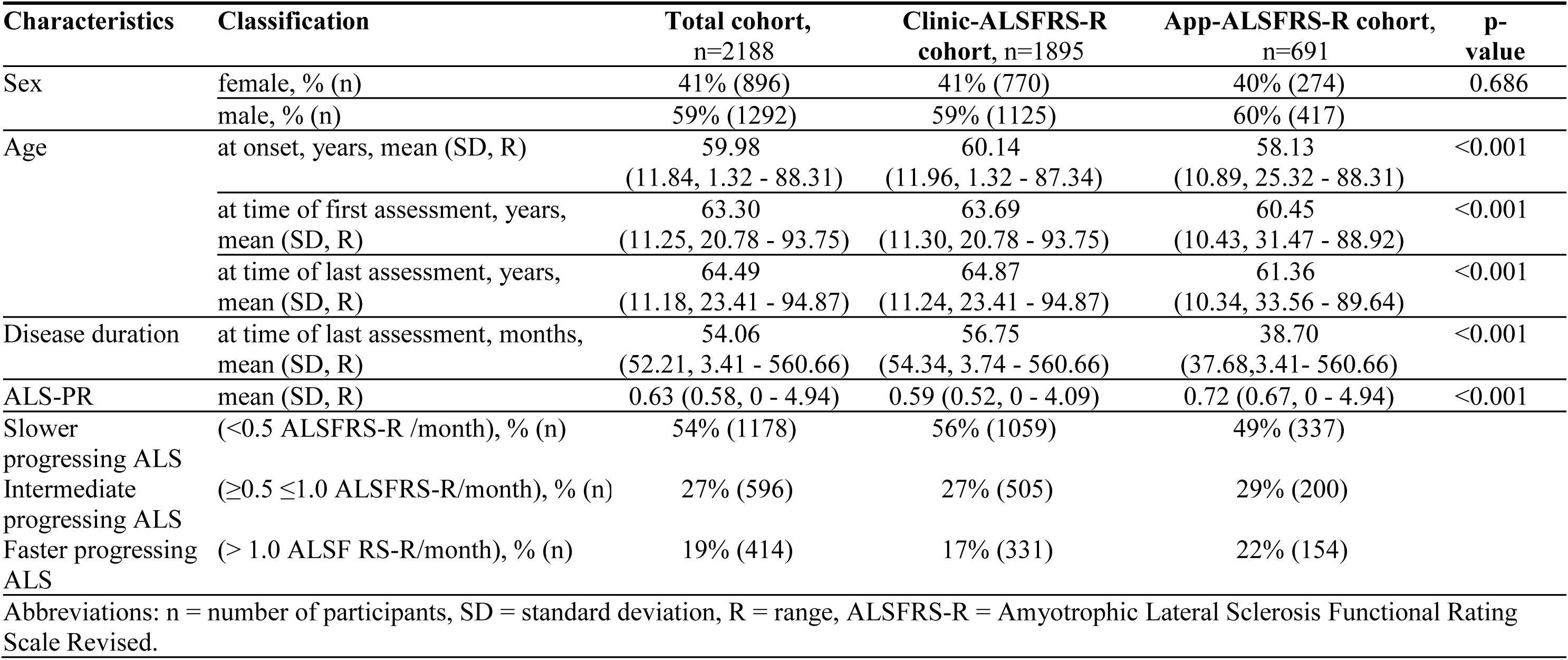
Demographic and clinical characteristics of participants.

#### ALSFRS-R

ALSFRS-R is a validated instrument to assess motor functions of the bulbar region, the extremities, the trunk including breathing abilities and the requirement for ventilatory support. It comprises 12 items with 5 rating options (0 to 4). The total range of the scale spans 0 (no function) to 48 scale points (full function). The ALSFRS-R was analyzed for all cohorts of this study.

#### ALSFRS-R-SE

The ALSFRS-R-SE includes instructions and explanations for each item, facilitating the assessment for healthcare professionals and patients (15). In this study, since May 9, 2022 the ALSFRS-R-SE replaced the ALSFRS-R on the ALS-App.

#### Classification of disease severity according to the ALSFRS-R total score

Disease severity was divided by four groups of disease severity according to ALSFRS-R total score: 48-37, 36-25, 24-13, 12-0 scale points.

#### ALS progression rate (ALSPR)

ALSPR was measured by the monthly change of ALSFRS-R scale points and calculated using the following formula: (48 minus ALSFRS-R total score divided by disease duration (months)).

#### Classification of ALSPR

ALSPR was divided in 3 groups: slower progressing ALS (<0.5 ALSFRS-R/month), intermediate progressing ALS (≥0.5 and ≤1.0 ALSFRS-R/month) and faster progressing ALS (> 1.0 ALSFRS-R/month).

#### Intrasubject variability (ISV) of ALSPR

ISV was assessed as variation of ALSPR: each patient with *n* ALSFRS-R assessments delivered *n* approximate ALSPR. To estimate the degree of variation, standard deviation (SD) or interquartile range (IQR) were considered. IQR was more robust against outliers and therefore chosen as the main comparative value.

### Statistical methods

Descriptive statistics were used for the statistical analysis (mean, standard deviation in ± and ranges).

#### Comparison of ISV of ALSPR

Null-hypothesis tests were applied to compare ISV of ALSPR for the App-ALSFRS-R and Clinic-ALSFRS-R cohorts, and to investigate whether or not significant differences could be found. For the total cohort a linear mixed model was applied, as some data met the criteria for dependency (combined ALSFRS-R cohort) and some patients were found solely in one cohort. The variation of ISV was determined as the target variable. Two-sample t-tests for paired samples were applicable to assess differences in ISV for the combined cohort.

#### Non-inferiority margin (δ)

To analyse non-inferiority a non-inferiority margin (δ) was determined. Results <δ would prove non-inferiority, as higher scores would indicate a higher ISV of ALSPR in App-ALSFRS-R data. Clinical reasoning and interpretation lead to defining δ as 0.15 IQR: a relatively high variability of the ALSFRS-R has been described before and was therefore anticipated (30). From clinical reasoning we determined, that a change in IQR of 0.15 ALSPR would still be acceptable.

#### Cohen’s d

Cohen’s d – or standardized mean difference – was determined to measure the effect size of the differences in ISV for the combined cohort. It was calculated as follows: The mean difference of IQR (mean IQR of App-ALSFRS-R minus mean IQR of clinic-ALSFRS-R) divided by SD for the statistical population. This allowed a standardized evaluation of the mean difference by relating it to standard deviation. The interpretation of the effect size varies in the literature. A commonly used interpretation is based on benchmarks: trivial effect (0.0-0.19), small effect (d=0.2), medium effect (d=0.5) and large effect (d=0.8) (31).

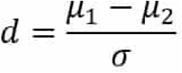

The data were analysed using ‘R’ Core Team version 4.4.0 (2024-04-24), R Foundation for Statistical Computing, Vienna, Austria.

## RESULTS

### Number of patients in cohorts

The total cohort encompassed 2188 ALS patients. The App-ALSFRS-R cohort consisted of 691 participants whereas the clinic-ALSFRS-R cohort included 1895 patients. 398 patients were found in the combined ALSFRS-R-cohort. **Figure 1**.

### Demographic and clinical characteristics

An overview of the demographic and clinical characteristics of the studied cohort is provided in **Table 1 and Table 2**. The clinical characteristics for the App-ALSFRS-R cohort and clinic-ALSFRS-R cohort were as follows: Mean age 60.45 (SD 10.43) and 63.69 (SD 11.3) years (p<0.001), mean disease duration 38.7 (SD 37.68) and 56.75 (SD 54.34) months (p<0.001), and mean ALSPR 0.72 and 0.59 (p<0.001), respectively. The App-ALSFRS-R cohort included more patients with faster progressing ALS compared the clinic-ALSFRS-R cohort (22%, n=154 vs 17%, n=331, respectively). Further differences were found between slower progressing ALS (49%, n=337 vs 56%, n=1059), and intermediate progressing ALS (29%, n=200 vs 27%, n=505) **Figure 2**.

**Figure 2.**
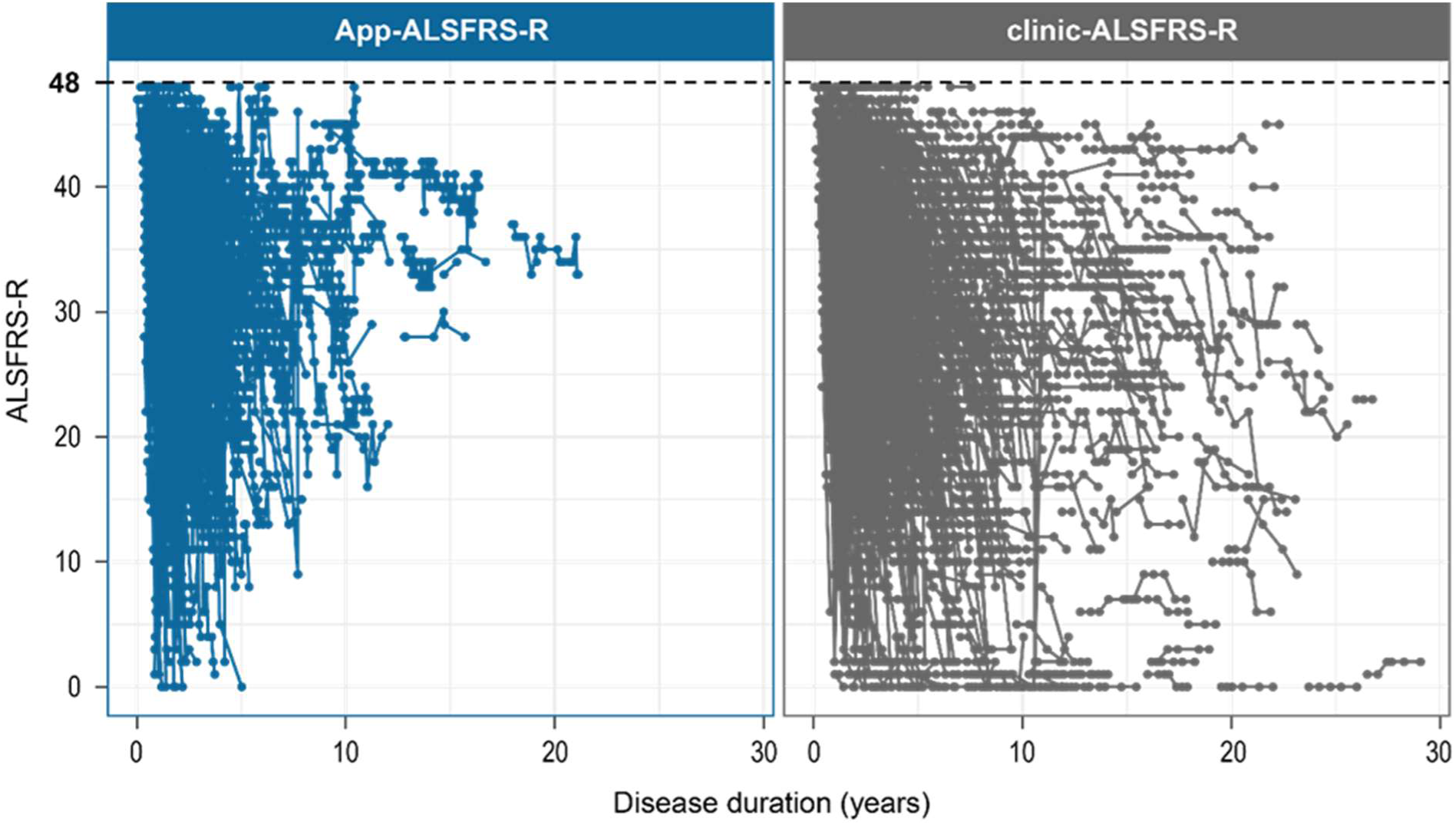
Overview of all App-ALSFRS-R and clinic-ALSFRS-R assessments based on the total score in comparison. Every dot represents one assessment. For depiction purposes, the disease duration was cut-off at 30 years. ALSFRS-R=Amyotrophic Lateral Sclerosis Functional Rating Scale revised, clinic-ALSFRS-R=ALSFRS-R assessed during clinic visits, App-ALSFRS-R=ALSFRS-R captured via self-assessment on patient’s smartphone.

**Table 2:**
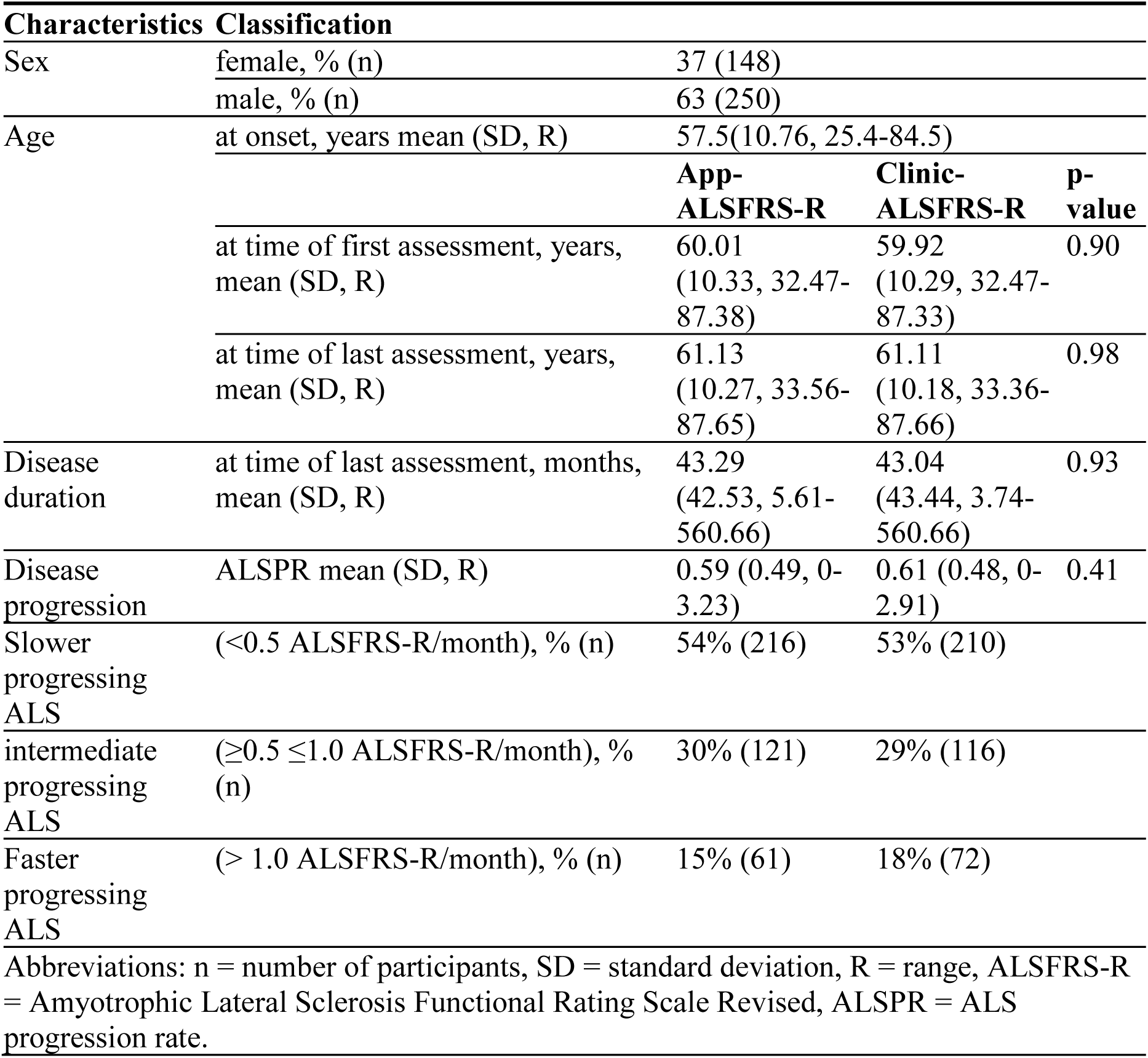
Demographic and clinical characteristics of the 398 patients in the combined cohort.

### Intrasubject variability (ISV) of ALSPR

The comparison of ISV of ALSPR for the total cohort was done by linear mixed model analysis. The IQR of the App-ALSFRS-R was reported at 0.171 (CI: 0.150, 0.191) and of clinic-ALSFRS-R 0.129 (CI: 0.119, 0.140). The difference did show statistical significance (p<0.001). **Table 3**.

**Table 3:** Comparison of intrasubject variability (ISV) of ALSPR.

| Classification | App mean<br>(95% CI) | Clinic mean<br>(95% CI) | p-value |
| --- | --- | --- | --- |
| **Total cohort** |  |  |  |
| IQR | 0.17<br>(0.15,0.91) | 0.13<br>(0.12,0.14) | <0.001 |
| **Combined cohort** |  |  |  |
| IQR | 0.12<br>(0.11, 0.14) | 0.12<br>(0.11, 0.14) | 0.242 |
| slower progressing ALS<br>(<0.5 ALSFRS-R/month) | 0.04<br>(0.03, 0.04) | 0.03<br>(0.03, 0.04) | 0.201 |
| intermediate progressing<br>ALS ($\geq 0.5 \leq 1.0$ ALSFRS-R/month), | 0.10<br>(0.08, 0.13) | 0.10<br>(0.08, 0.12) | 0.733 |
| faster progressing ALS<br>($> 1.0$ ALSFRS-R/month) | 0.25<br>(0.18, 0.33) | 0.22<br>(0.16, 0.29) | 0.419 |
| Abbreviations: ALSPR = Amyotrophic lateral sclerosis progression rate, CI = confidence interval, IQR = interquartile range |  |  |  |

The combined ALSFRS-R cohort was analysed separately. **Table 3**. The mean ISV of ALSPR in the App-ALSFRS-R cohort was 0.12 (IQR, CI: 0.11,0.14), and in the clinic-ALS-FRS-R at 0.12 alike (IQR, CI: 0.11,0.14). A two-sample t-test for paired samples confirmed no significant difference in the IQR of ISV in both cohorts (t-test p-value: 0.242). The upper limit of CI of App-ALSFRS-R was below the predefined non-inferiority margin, demonstrating non-inferiority.

The mean difference in ISV of the ALSPR between App-ALSFRS-R and clinic-ALSFRS-R was 0 (IQR, SD 0.13). To further asses the effect size of this difference Coheńs d was determined. Cohen’s d was 0.06 (IQR, CI: −0.14, 0.26), which is interpreted as a trivial effect of the difference in ISV (31). **Figure 3**.

**Figure 3.**
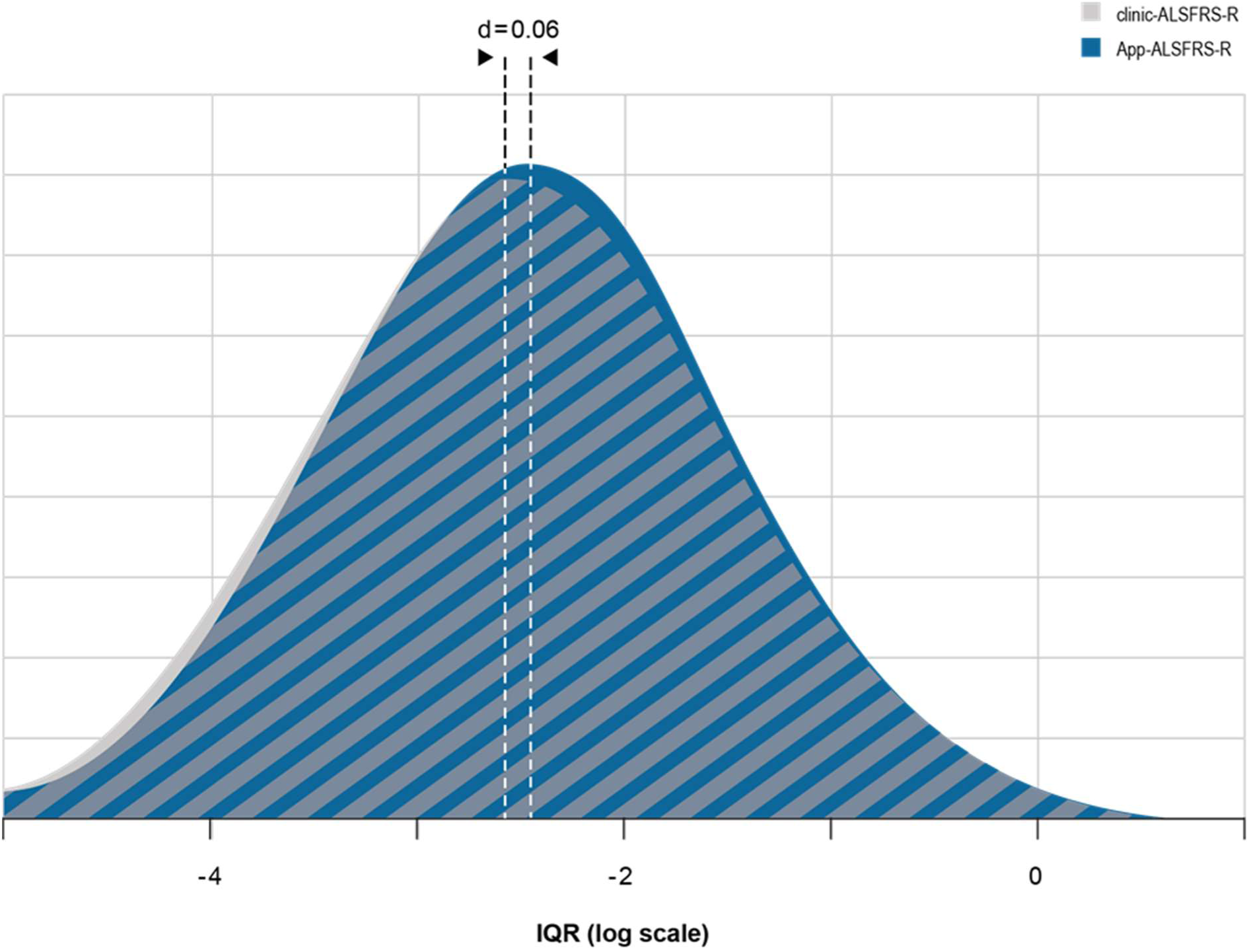
Visualization of Coheńs d. The effect size of the standardized mean difference in intrasubject variability of App-ALSFRS-R and clinic-ALSFRS-R, assessed as interquartile range (IQR), was determined. The result of 0.06 can be interpreted as a trivial effect, meaning the IQR showed an overlap 97.6% for the compared methods. d=Coheńs d, ALSFRS-R=Amyotrophic Lateral Sclerosis Functional Rating Scale revised, clinic-ALSFRS-R=ALSFRS-R assessed during clinic visits, App-ALSFRS-R=ALSFRS-R captured via self-assessment on patient’s smartphone

### ISV of ALSPR in groups of ALSPR

The ISV of three groups of ALSPR of the combined cohort were compared in a subgroup analysis, to assess a possible impact of different ALSPR on ISV. Although statistically not significant (p=0.141), ISV of ALSPR of faster progressing ALS demonstrated a trend towards higher ISV in both, clinic-ALSFRS-R (IQR 0.217, 95% CI: 0.161, 0.292) and App-ALSFRS-R (IQR 0.246, 95% CI: 0.183,0.332; p=0.419) compared to slower progressing ALS with 0.035 (IQR, CI: 0.030, 0.041; p=0.201) and 0.031 (IQR, CI: 0.027, 0.037) as well as intermediate progressing ALS with 0.101 (IQR, CI: 0.080, 0.128) and 0.097 (IQR, CI: 0.077, 0.123; p=0.733), respectively. Significant differences in ISV between clinic-ALSFRS-R and App-ALSFRS-R were not found for any of the studied groups of ALSPR **(Figure 4 and 5a)**.

**Figure 4.**
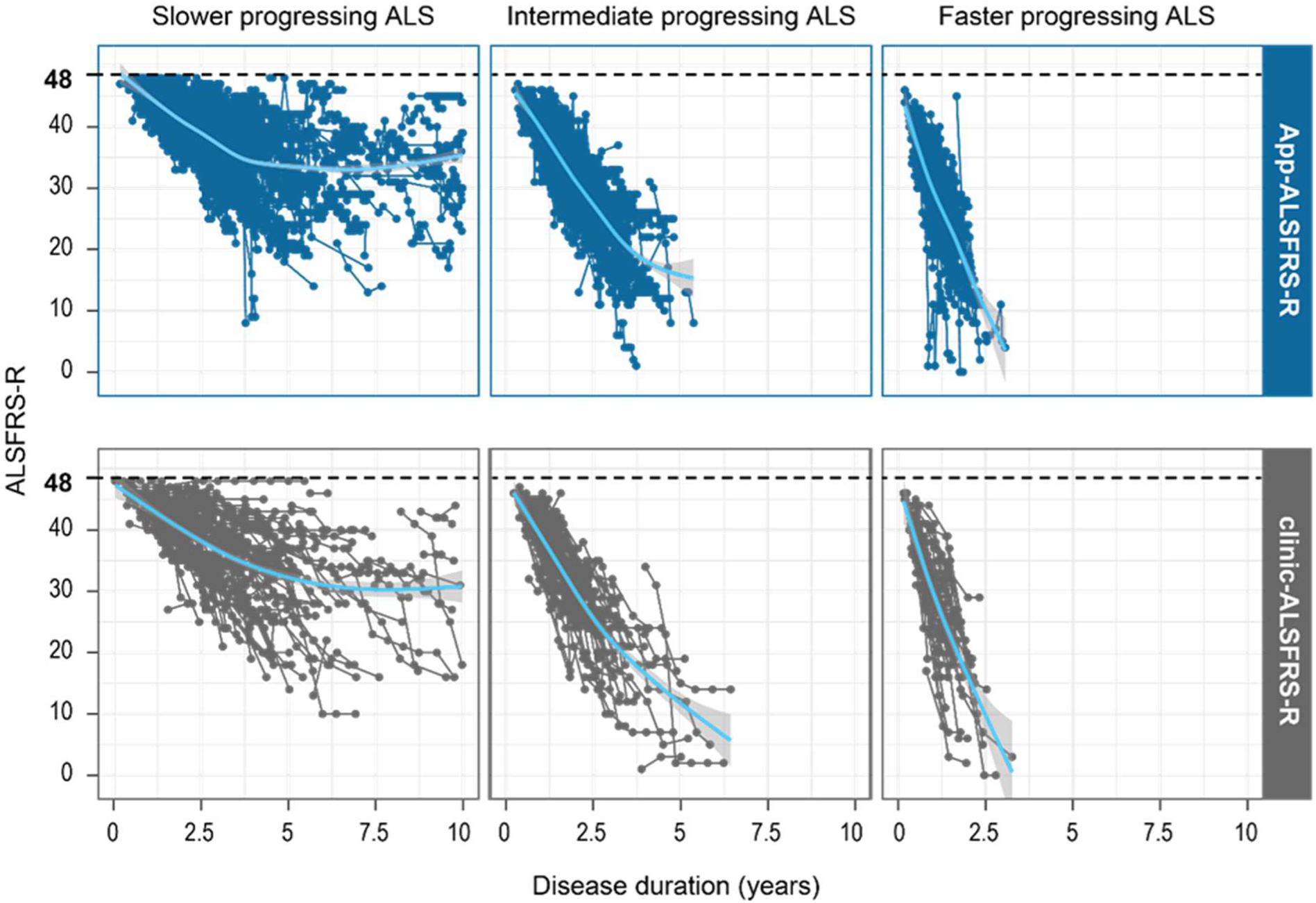
Comparison of clinic and remote assessment of the ALSFRS-R based on the total score. Overview of the assessments of App-ALS-FRS-R and clinic-ALS-FRS-R as analyzed in 3 groups of ALS progression rate (ALSPR), respectively: slower progressing ALS (<0.5 ALSFRS-R/month), intermediate progressing ALS (≥0.5 and ≤1.0 ALSFRS-R/month) and faster progressing ALS (> 1.0 ALSFRS-R/month). Every dot represents one assessment. The mean progression is shown in the blue graph and the shadow represents its variation. ALSFRS-R=Amyotrophic Lateral Sclerosis Functional Rating Scale revised, clinic-ALSFRS-R=ALSFRS-R assessed during clinic visits, App-ALSFRS-R=ALSFRS-R captured through self-assessment on patient’s smartphone

**Figure 5.**
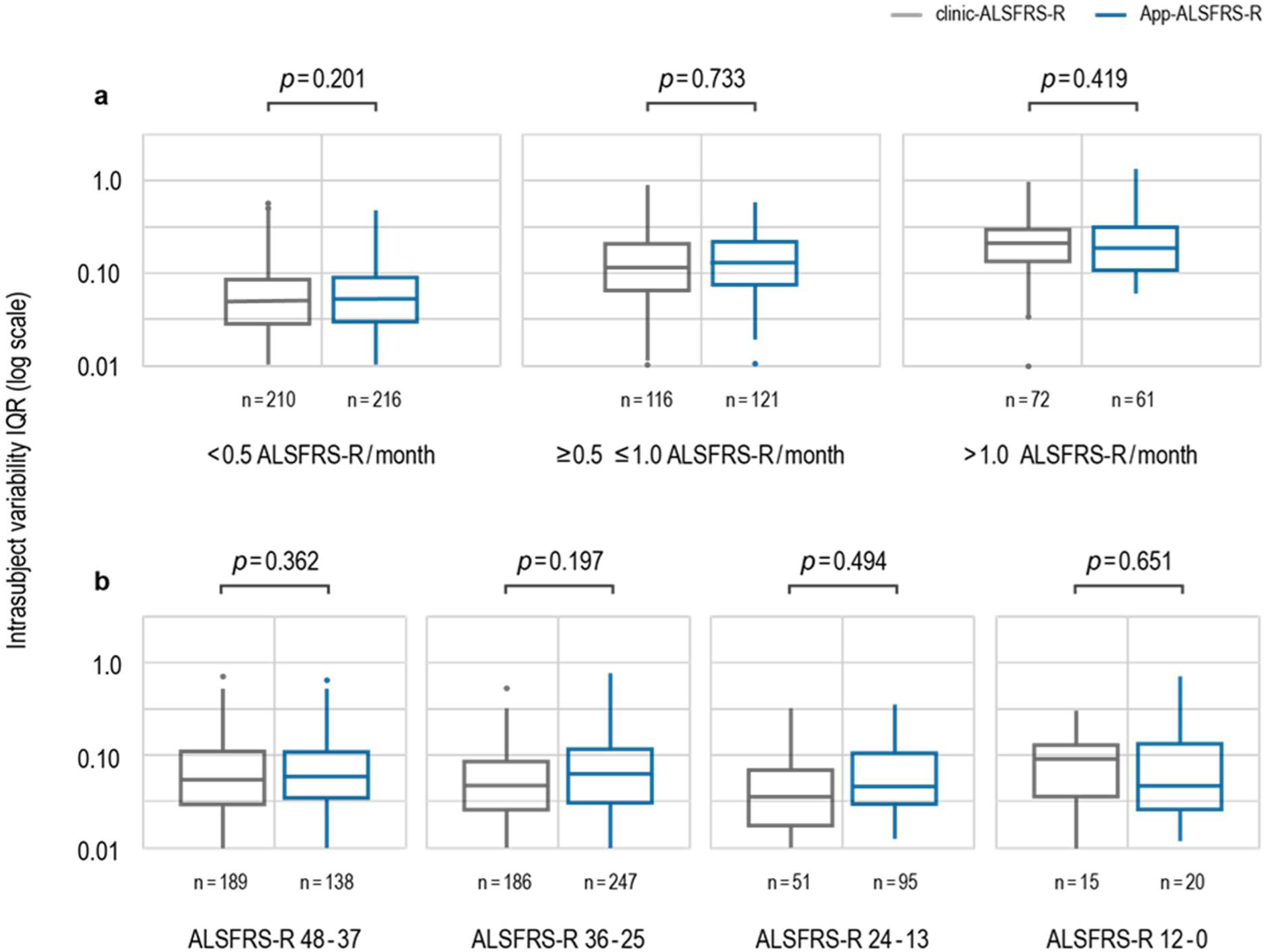
Comparison of clinic and remote assessment of the ALSFRS-R based on the ALS progression rate (ALSPR). The intrasubject variability (ISV) of the ALSPR – as calculated from the ALSFRS-R assessment – was investigated to compare the App-ALSFRS-R and clinic-ALSFRS-R. The analysis was stratified for a) three groups of ALSPR: slower, intermediate and faster progressing ALS based on the ALSPR and b) four groups of disease severity according to ALSFRS-R total score. The numbers for each group exceed the total number of patients, as individuals contributed data in several groups of disease severity or ALSPR, respectively. n=number of patients, IQR=Interquartile range, ALSFRS-R=Amyotrophic Lateral Sclerosis Functional Rating Scale revised, clinic-ALSFRS-R=ALSFRS-R assessed during clinic visits, App-ALSFRS-R=ALSFRS-R captured via self-assessment on patient’s smartphone

### ISV of ALSPR in groups of disease severity according to ALSFRS-R total score

The comparison between App-ALSFRS-R and clinic-ALSFRS-R did not show significant differences: ALSFRS-R 48-37: 0.051 (IQR, CI 0.043, 0.062) and 0.047 (IQR, CI 0.039, 0.057; p=0.362); ALSFRS-R 36-25: 0.042 (IQR, CI 0.035, 0.051) and 0.037 (IQR, CI 0.031, 0.045; p=0.197); ALSFRS-R 24-13: 0.033 (IQR, CI 0.022, 0.050) and 0.028 (IQR, CI 0.019, 0.043; p=0.494); ALSFRS-R 12-0: 0.053 (IQR, CI 0.015, 0.189) and 0.072 (IQR, CI 0.020, 0.257; p=0.651), respectively **(Figure 5b).**

## DISCUSSION

The technological and methodological means for remote digital assessment of ALSFRS-R is on the rise for several years (7). This trend has been supported by previous reports on a strong correlation of on-site and online assessments (6,32,33). This study focussed specifically on ALSFRS-R data capture on the patient’s smartphones and the open research question of its non-inferiority when compared to clinic assessment.

Patientś readiness to perform remote assessment of ALSFRS-R was shown before (7) and confirmed in this study. The large number of 691 ALS patients using the ALS-App, who completed at least two assessments of the ALSFRS-R, facture in the strength of this study. Remarkably, 293 patients remotely provided App-ALSFRS-R data, although they had only realized one clinic visit. Also, the large number of App-ALSFRS-R were generated – in addition to the conventional source of ALSFRS-R data. Both findings contribute to the notion, that digital assessments increase the data density between visits, can potentially fill data gaps of missing visits or might extend information on the disease course, even when clinic visits are not possible anymore.

Previous studies consistently reported a higher ALSFRS-R total score in digital assessments when compared to clinic ALSFRS-R data capture (13,32–34). Even though we did not explicitly investigate the differences in total scores, this study confirmed those findings. In line with previous research on digital assessment of the ALSFRS-R, participants of remote assessment were younger and earlier in the course of ALS (7). This difference may be explained by a selection bias related to technical barriers as well as time efforts of using of digital and telemedicine devices (7,17,33–35). Thus, the ALS-App was commonly offered at the patient’s first visit in the respective study centres. Patients with a very long disease course were possibly not considered by the recruiters. Furthermore, it is conceivable that patients in the earlier course of ALS were overrepresented as the ALS-App may have received more attention in patients with newly diagnosed ALS. Furthermore, the findings might point to barriers for patients with lower motor functional capacities. Future research must aim to apply patient-centred services, technical support and app design to warrant patient’s access to digital assessment in all phases on the disease. It is worth mentioning though, that compared to other epidemiological data on ALS cohorts, the mean disease duration in this cohort was relatively long (36). A previously described difference in gender distribution vanished and female patients participated in digital assessment proportionally equal to male patients.

Patients using the ALS-App showed a more aggressive disease progression, as the mean ALSPR was higher, than in the clinic cohort. Correspondingly, faster progressing ALS was overrepresented in the ALS-App cohort. This finding suggests that self-assessment of ALSFRS-R on a mobile device is feasible for patients even with faster progressing ALS and of importance when considering remote digital assessments in clinical trial settings. Also, in clinical practice, as a faster progression can make clinic visits more burdensome or even impossible, this observation supports the feasibility of the App-use in a wide clinical spectrum of ALS. The differences between faster and slower progressing ALS among ALS-App users can be discussed from a different angle – patients with a more aggressive disease course might perceive more relevance and need to report on the progression of ALS than patients with a slower disease course. At the same time, patients with slower disease and less changes in the ALSFRS-R over time might be less motivated to frequently and continuously provide self-ratings.

Inter- and intrarater variability of ALSFRS-R and ALSPR was in the focus of this investigation. Previous research showed contradicting results of a higher, but mostly lower variability of remote-self assessments (6,13,33,35). The clinic ALSFRS-R cohort was subject of both, inter- and intrarater variability, as the assessment during clinic visits was performed by variable raters that might have changed from visit to visit. In principle, in the ALS-App cohort only intrarater variability was assumed. However, it cannot certainly be excluded that some patients shared login data and authorized relatives to perform the assessment. In this unwanted constellation interrater variability was caused, which belongs to the limitations of the study.

The comparison between the clinic and app cohort as well as the investigation of non-inferiority was performed based on ALSPR, but not ALSFRS-R total score. When predicting the course of ALS in clinical practice and trials, ALSPR, commonly named ‘slop’ or ‘delta ALSFRS-R’, is more informative than the total score (23). This study revealed no difference of ISV of ALSPR between App-ALSFRS-R and clinic ALSFRS-R and CI below a predefined non-inferiority margin for the statistically robust combined cohort. Furthermore, the data indicated a trivial (as assessed by Coheńs d) difference. Overall, this study proved non-inferiority of App-ALSFRS-R compared to clinic ALSFRS-R. When we stratified the combined cohort for classes of ALSPR we found, that the ISV increased from slower to faster progressing ALSPR. Although this was not significant, a higher variability must be expected, when faster progressing ALSPR is investigated.

This may also explain the lower ALSPR-ISV of the total clinic-ALSFRS-R-cohort in comparison to the total App-ALSFRS-R-cohort, as the latter was characterized by a higher mean ALSPR. Although reasons for differences in the total cohort can be various: e.g. the methodologically caused dependency on the onset date, the longer disease duration in the total clinic cohort compared to the total app cohort and overall different sample sizes.

An important limitation in the presented method is the dependency of the ALSPR on the onset date which is based on the patient’s recollection of the time of the start of dysarthria, dysphagia, limb paresis or (rarely) hypoventilation. The training of evaluators – and even more importantly of the patients – is crucial to define the onset date in a consented and therefore, harmonized manner.

The possible impact of disease severity, as measured by the total score of ALSFRS-R, was studied in the combined cohort and did not show significant difference in ISV of ALSPR. This observation underscored methodological feasibility of ALS-App use during the complete course of disease, including very progressed phases of ALS. On this basis, patients with progressed ALS and greater barriers for clinic visits can be offered to use the ALS-App for digital assessment of ALSFRS-R, mainly to gain functional information and to support care related decision-making from remote. This conclusion comes with some limitation as a non-significant difference in ISV of ALSPR was found in the group lowest motor function (0-12 points). In this stage of the disease, changes in respiratory items (items 10-12) become most relevant which is known to be subject of greatest variability (27). This emphasizes the need for training of evaluators, when assessing the ALSFRS-R and the potential benefits of self-explanatory ALSFRS-R-SE.

In summary, this observational study supported the concept of remote digital assessment of ALSFRS-R and proved non-inferiority of ALSFRS-R data being captured on the patient’s smartphone – compared to the clinic assessments. Our findings suggest that app assessments can increase ALSFRS-R data density between clinic visits, might fill data gaps of missing onsite visits or allow the remote assessment of the ALSFRS-R in progressed phases of ALS, when clinic visits are burdensome and in protracted intervals.

## Acknowledgements

The authors thank all the patients who gave their effort and valuable time to participate in this study. The authors wish to thank the Boris Canessa ALS Stiftung (Düsseldorf) for co-funding this work and continuous support.

## Declaration of interest statement

TM has received grants, personal fees, non-financial support and research support from AL-S Pharma, Amylyx, Cytokinetics, Ferrer, Mitsubishi Tanabe, Sanofi, Orphazyme, and served on the advisory boards of Amylyx, Biogen, and ITF Pharma outside of the submitted work. TM and CM are founders and shareholders of the Ambulanzpartner Soziotechnologie APST GmbH, which makes the mobile application “ALS-App”.TG has received personal fees from ITF Pharma and served on the advisory boards of Amylyx and ITF Pharma outside of the submitted work. SP has received speaker fees, non-financial support and research support from Biogen, Roche, ALS Pharma, Amylyx, Cytokinetics, Ferrer, ITF Pharma, and Sanofi and served on advisory boards of Amylyx, Biogen, Roche, Zambon and ITF Pharma outside of the submitted work. PW has served on advisory boards of Biogen, ITF Pharma and Novartis outside of the submitted work. RG has received grants, personal fees, non-financial support and research support from Biogen and served on the advisory boards of Biogen, Roche, and ITF Pharma outside of the submitted work. MV received travel expenses and non-financial support from ITF Pharma outside of the submitted work. AM has received personal fees, non-financial support, and research support from ITF Pharma and Zambon outside the submitted work. PK received consulting fees from Biogen. PL reports grants from the Bundesministerium für Bildung und Forschung and the Deutsche Forschungsgemeinschaft; consulting fees from AbbVie, Amylyx, Bial, Desitin, ITF Pharma, Novartis, Stadapharm, Raya Therapeutic, Woolsey Pharmaceuticals, and Zambon; and is co-inventor on a patent for the use of fasudil in amyotrophic lateral sclerosis (EP 2825175 B1, US 9.980,972 B2), outside of the scope of the submitted work. The other authors declare no conflicts of interest.

## Authoŕs contribution

TM, LS and AM designed and conceptualized the study, analyzed and interpreted the data, and drafted the manuscript for intellectual content. BW had a major role in data collection and preparation of data. TG, UW, DK, RS, AR, JG, SP, MB, PW, SB, RG, PL, JCK, PB, JHW, JD, YK, IC, MV, JN, PS, PK, SSP and CM had a major role in data acquisition and revised the manuscript for intellectual content.

## Data availability statement

The data that support the findings of this study are available from the corresponding author upon reasonable request.

## Funding

This work was supported by the Boris Canessa ALS Stiftung (Düsseldorf).

## Notes

### Competing Interest Statement

TM has received grants, personal fees, nonfinancial support and research support from ALS Pharma, Amylyx, Cytokinetics, Ferrer, Mitsubishi Tanabe, Sanofi, Orphazyme, and served on the advisory boards of Amylyx, Biogen, and ITF Pharma outside of the submitted work. TM and CM are founders and shareholders of the Ambulanzpartner Soziotechnologie APST GmbH, which makes the mobile application ALS App.TG has received personal fees from ITF Pharma and served on the advisory boards of Amylyx and ITF Pharma outside of the submitted work. SP has received speaker fees, nonfinancial support and research support from Biogen, Roche, ALS Pharma, Amylyx, Cytokinetics, Ferrer, ITF Pharma, and Sanofi and served on advisory boards of Amylyx, Biogen, Roche, Zambon and ITF Pharma outside of the submitted work. PW has served on advisory boards of Biogen, ITF Pharma and Novartis outside of the submitted work. RG has received grants, personal fees, nonfinancial support and research support from Biogen and served on the advisory boards of Biogen, Roche, and ITF Pharma outside of the submitted work. MV received travel expenses and nonfinancial support from ITF Pharma outside of the submitted work. AM has received personal fees, nonfinancial support, and research support from ITF Pharma and Zambon outside the submitted work. PK received consulting fees from Biogen. PL reports grants from the Bundesministerium fuer Bildung und Forschung and the Deutsche Forschungsgemeinschaft; consulting fees from AbbVie, Amylyx, Bial,
Desitin, ITF Pharma, Novartis, Stadapharm, Raya Therapeutic, Woolsey
Pharmaceuticals, and Zambon; and is coinventor on a patent for the use of fasudil in amyotrophic lateral sclerosis (EP 2825175 B1, US 9.980,972 B2), outside of the scope of the submitted work. The other authors declare no conflicts of interest.

### Funding Statement

This work was supported by the Boris Canessa ALS Stiftung (Duesseldorf).

### Author Declarations

The study protocol was approved by the Medical Ethics Committee of Charite Universitaetsmedizin Berlin, Germany under number EA1/219/15. A signed patient information and informed consent form was obtained from all the participating patients.

